# Effect of moderate altitude with and without nocturnal oxygen therapy on the next-day cerebrovascular function in patients with chronic obstructive pulmonary disease – A randomized, cross-over trial at 2048 m

**DOI:** 10.1101/2025.01.27.25321003

**Authors:** Dominic Gilliand, Tsogyal D. Latshang, Sayaka S. Aeschbacher, Fabienne Huber, Deborah Flueck, Mona Lichtblau, Stefanie Ulrich, Elisabeth D. Hasler, Philipp M. Scheiwiller, Julian Müller, Silvia Ulrich, Konrad E. Bloch, Michael Furian

## Abstract

**Background:** This study investigated whether nocturnal oxygen therapy (NOT) improves next-day cerebrovascular function in lowlanders with COPD staying at moderate altitude.

**Methods:** This randomized, placebo-controlled crossover trial was performed in stable patients with moderate to severe COPD (FEV_1_/FVC <0.7 and FEV_1_ 30-80%predicted), living <800m and pulse oximetry (SpO_2_) ≥92%. Patients underwent assessments at 490m and during 2 stays of 2 days at 2048m while NOT or placebo (each at 3L min^−1^ through nasal cannula) were applied according to a randomized cross-over design. At both altitudes, SpO_2_, cerebral tissue oxygenation (CTO, near-infrared spectroscopy), mean arterial blood pressure (MAP, finger plethysmography) and middle cerebral artery systolic peak blood flow velocity (sMCAv, transcranial Doppler ultrasound) were assessed while patients were *(0)* quietly breathing FiO_2_ 0.21; *(i)* quietly breathing FiO_2_ 1.0, *(ii)* voluntarily hyperventilating, *(iii)* voluntarily hyperventilating under FiO_2_ 1.0, and *(iv)* during head-up tilting. Indices of cerebrovascular responsiveness to changes in blood gases and blood pressure were computed.

**Results:** A total of 18 patients (8 women aged mean±SD 65±5y, FEV_1_ 54.7±13.9%predicted) were analyzed. At 2048m under placebo, patients became hypoxemic, mean±SE SpO_2_ 90.3±0.4% vs. 93.7±0.4% at 490m, while MAP, CTO and sMCAv remained unchanged compared to 490m. All ventilatory maneuvers at 2048 m induced greater increases in SpO_2_ compared to 490m while changes in MAP, CTO and sMCAv were similar. Head-up tilting induced a similar blood pressure fall at 2048m compared to 490m, whereas cerebral blood flow velocity changed less in response to systemic hypotension (mean±SE ΔsMCAv/ΔMAP 0.9±0.3 vs. 2.3±0.4cm s^−1^ mmHg^−1^) at 2048m. No alteration in cerebrovascular function as a treatment effect of NOT was observed in either maneuver.

**Conclusion:** This randomized clinical trial in moderate-to-severe COPD patients ascending to 2048m showed that moderate daytime systemic hypoxemia does not translate to cerebral hypoxia nor cerebrovascular autoregulatory impairments while at rest or under ventilatory or orthostatic challenges.

## INTRODUCTION

Chronic obstructive pulmonary disease (COPD) is a systemic disease characterized by reduced lung function and airflow obstruction and is one of the world’s leading causes of death. Given the high prevalence of COPD of 8 – 15 % of the adult world population and recent improvements in symptom-mediated treatments, it is assumed that many COPD patients are among individuals living at low but ascending to high altitude for work or recreational activities (1). Acute exposure to hypobaric hypoxia at higher elevations may pose a challenge for an impaired respiratory system with less ventilatory reserves (2). Previous studies have shown that some patients with stable moderate to severe COPD develop altitude-related illnesses (3–5), experience exercise intolerance, disturbed sleep and disturbed breathing patterns at altitude (6–9). Moreover, it has been shown that already at moderate altitude (2048 m), cerebrovascular autoregulation was unable to protect the brain from cerebral oxygen desaturations while sleeping and under physical exertion (3, 9). Whether cerebrovascular homeostasis and responsiveness in COPD is impaired under quiet daytime conditions at moderate altitude has not been studied.

In a randomized clinical trial investigating the preventive effect of nocturnal oxygen therapy (NOT) in COPD at moderate altitude, mixed effects were observed. Tan and colleagues (3) studied the effect of NOT on altitude-associated periodic breathing during sleep at 2048 m. They found an improvement in nocturnal arterial and cerebral oxygenation and stabilized breathing patterns at night compared to placebo. The participants furthermore had significantly improved subjective sleep quality and experienced less high altitude-related illnesses. Within the same trial, Meszaros et al. have shown that NOT mitigated altitude-induced rise in systolic BP and elevated baroreflex-sensitivity compared to placebo at altitude (10). Whether NOT improves cerebrovascular function the next morning at physical rest, remains to be elucidated.

Therefore, the aim of this study was to investigate altitude-related changes in the cerebrovascular homeostasis and responsiveness under resting conditions and to elucidate the effects of NOT in patients with COPD staying overnight at moderate altitude.

## METHODS

### Study design and intervention

This study is part of a randomized, placebo-controlled crossover trial performed at the University Hospital of Zurich (490 m) and at St. Moritz (2048 m) to evaluate the effect of NOT for COPD patients on nocturnal hypoxemia, altitude-associated periodic breathing, and altitude-related illnesses. Outcomes above have been published previously (3). The main trial has been registered at clinicaltrials.gov with the identifier NCT02150590. All participants gave informed written consent. The trial has been approved by the institutional ethics committee (EK-2013-0088). Other findings of this study have been published elsewhere (3, 9–12), whereas the current findings related to the cerebrovascular homeostasis and responsiveness have not been published.

Participants performed baseline measurements at 490 m and during two sojourns at 2048 m for a period of two days and two nights per stay with a washout period of two weeks in between. At 2048 m, participants were randomly allocated to receive either NOT (100% oxygen, FiO_2_ 1.0) or placebo (ambient air, FiO_2_ 0.21) at 3 L min^−1^ flow rate via nasal cannula during the night and were blinded to this intervention. Transfers between 490 m and 2048 m were made by train and car within 3 hours. Usual medication was continued during the trial.

### Participants

Eligible patients of all genders, diagnosed with moderate to severe COPD according to the GOLD criteria (with FEV_1_/FVC <0.7 and FEV_1_ between 30 - 80%), aged 18 - 75 years and living <800 m, were invited to participate in this study. Criteria for exclusion were past exacerbation of COPD within 4 months before the trial, oxygen saturation <92% at 490 m, any uncontrolled cardiovascular disease, previous altitude intolerance or altitude exposure >1500 m for >2 days within the last 4 weeks before the trial. During the trial, patients were withdrawn from the study for safety reasons if they experienced one of the following predefined altitude-related adverse health effects; systolic or diastolic blood pressure >200 mmHg or >110 mmHg, respectively, SpO_2_ <75% at rest for >30 minutes, acute chest pain at rest, acute mountain sickness, COPD exacerbation or any other condition requiring medical treatment or relocation to low altitude.

### Maneuvers to assess cerebrovascular function

At 490 m and on the second day at 2048 m, patients underwent cerebrovascular testing in calm supine condition while performing instructed breathing maneuvers followed by a supine-to-60° head-up tilt. The breathing maneuvers aimed to produce different states of arterial partial pressure of CO_2_ and O_2_ (PaCO_2_ and PaO_2_). Cerebral blood flow (CBF) was assessed during the following conditions with the breathing maneuvers intended to induce a respective intensification of cerebral vasoconstriction: (*0*) poikilocapnic hypoxia with quiet room air breathing, *(i)* poikilocapnic hyperoxia with quiet oxygen breathing (FiO_2_ 1.0), *(ii)* hypocapnic normoxia with room air hyperventilation, and (*iii)* hypocapnic hyperoxia with oxygen breathing hyperventilation. In a specified sequence cycle, participants were asked to carry out 10 minutes of quiet breathing, followed by an episode of hyperventilation until a plateau of the end-tidal partial pressure of CO_2_ (PetCO_2_) was reached for at least 15 seconds. This sequence was carried out twice in a row while either placebo (ambient air FiO_2_ 0.21) or oxygen (FiO_2_ 1.0) was administered to the participants via face mask with reservoir bag. The order of placebo or oxygen administration for the breathing maneuvers was randomized. After the breathing maneuvers, a washout-period of at least 10 minutes was awaited, for normalization of all parameters. In the end, *(iv)* a head-up tilt was applied by inclining the bed to 60° (ergoline GmbH, Bitz, Germany) for 3 minutes to provoke a rapid reduction of mean arterial blood pressure (MAP). All measurements were performed in the morning.

### Measurements

During all maneuvers, continuous variable measurements were performed. Peak systolic blood flow velocity of the middle cerebral arteries (sMCAv) was measured by transcranial Doppler ultrasound (TOCM, Multigon Industries, New York, USA) with 2-MHz probes placed bilaterally at the temporal windows. The position and settings were identical at each session within a patient. Beat-by-beat blood pressure and heart rate were measured non-invasively with the finger-cuff technique (Finapres Midi, FMS, The Netherlands) with the finger positioned at heart level. Systolic and diastolic blood pressure values of the finger where calibrated by the mean of 3 brachialis sphygmomanometer measurements. Breath-by-breath PetCO_2_ was assessed by capnography (Capnocheck Sleep; Smiths Medical PM Inc., Waukesha, WI, United States). Additional measurements included pulse oximetry (SpO_2_) and cerebral near-infrared spectroscopy (NIRO 200NX device, Hamamatsu, Japan) assessing cerebral tissue oxygenation (CTO), oxygenated-(O_2_Hb) and deoxygenated hemoglobin concentration (HHb) as surrogates for changes in local cerebral oxygenation and total hemoglobin concentration (totHb) as index of local cerebral blood volume.

Averages of all parameters were calculated using data that were measured during predefined time periods. Measurements of the quiet breathing maneuvers were conducted during the last 3 minutes of the period at rest with a 30 second gap before the start of hyperventilation, and the last 15 seconds of the PetCO_2_ plateau during the hyperventilation maneuvers were analyzed. For the head-up tilt, the corresponding nadir of the sMCAv and MAP curve were considered to pronouncedly demonstrate the effect of the maneuver from baseline.

### Outcome calculation

Cerebrovascular responsiveness to blood gases was assessed in change of systolic MCAv per unit change of PetCO_2_ (ΔsMCAv/ΔPetCO2 (cm s^−1^ mmHg^−1^)) and SpO_2_ (ΔsMCAv/ΔSpO_2_ (cm s^−1^ %^−1^)) (13).

During the time period of the head-up tilt, dynamic cerebral autoregulation was quantified by the total unit reduction of sMCAv per unit reduction of MAP (ΔsMCAv/ΔMAP (cm s^−1^ mmHg^−1^)). The percent reduction of MCAv per unit reduction of MAP (%ΔsMCAv/ΔMAP (% mmHg^−1^)) and in percent reduction of sMCAv per percent reduction of MAP (%ΔsMCAv/%ΔMAP (% %^−1^)) with greater values indicating more impaired autoregulation (14). Furthermore, the cerebrovascular conductance index (CVCi = sMCAv/MAP (cm s^−1^ mmHg^−1^)), and its reciprocal the cerebrovascular resistance index (CVRi = MAP/sMCAv) was calculated for each maneuver assuming a stable vessel diameter.

### Outcomes

The main outcomes are the variables reflecting changes in sMCAv in response to PetCO_2_, SpO_2_ and MAP alterations as described in the section above. Secondary outcomes include the CVCi and CVRi, and the changes of parameters obtained from near-infrared spectroscopy for each maneuver, respectively.

### Statistical analysis

The data are summarized as mean and SD. To assess the effect of an ascent from 490 m to 2048 m and the effect of NOT compared to placebo on cerebrovascular responsiveness, a mixed linear regression model was conducted using patients as random effects and altitude (490 m, 2048 m), nocturnal intervention (NOT, placebo) and cerebrovascular challenges (*i to v*) and their interaction as fixed effects. P<0.05 reflect statistically significant differences. Data analysis was performed in the per-protocol population, defined as patients visiting undergoing all scheduled assessments and both measurements for MAP and sMCAv were successfully performed within the required time frame for at least one of four maneuvers. Statistical analysis was performed in STATA 15.

## RESULTS

### Study flow and patients’ characteristics

A total of 32 patients with moderate to severe COPD were randomized for this study. At 2048 m, four patients experienced severe nocturnal hypoxemia defined as SpO_2_ <75 % for <30 minutes, 2 patients developed an exacerbation of their COPD, 2 patients wished to be relocated to low altitude for feeling ill, 1 patient experienced a panic attack and 1 patient had non-sustained ventricular tachycardia (3). Furthermore, 4 patients could not be included in the final analysis due to technical failures. Therefore, 18 patients finished the cross-over trial and were included in the final analysis (**Figure 1**). The study participants comprised of 8 (44 %) women and 10 (56 %) men aged 65 ± 5 years with a mean FEV_1_ of 54.7 ± 13.9 % predicted. In **Table 1**, the patient’s characteristics are summarized.

**Figure 1.**
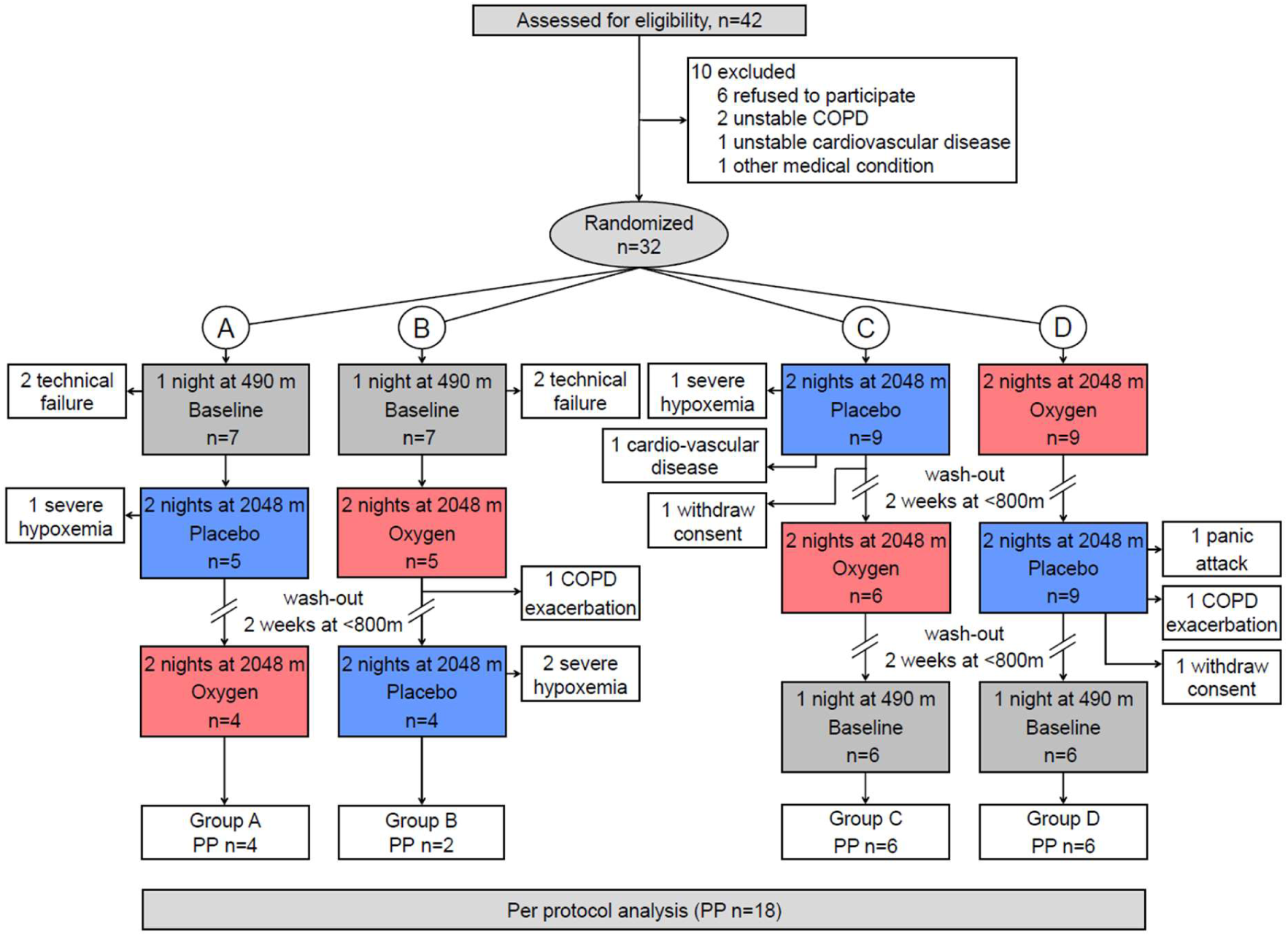
Study flow chart. COPD, chronic obstructive pulmonary disease; PP, per-protocol.

**Table 1.** Patient characteristics of the per-protocol population.

| *Lowland living participants with COPD* |  |
| --- | --- |
| **N (women), n** | 18 (8) |
| **Age, years** | 65 ± 5 |
| **Height, cm** | 169 ± 0.1 |
| **Weight, kg** | 73.5 ± 12 |
| **BMI, kg/m<sup>2</sup>** | 25.8 ± 4.0 |
| **Smoking status** |  |
| Current, n (%) | 4 (22.2) |
| Former, n (%) | 14 (77.8) |
| **Smoking, pack-years** | 45.2 ± 31.2 |
| **FEV<sub>1</sub>, % predicted** | 54.7 ± 13.9 |
| **FVC, % predicted** | 80.3 ± 17 |
| **FEV<sub>1</sub>/FVC, %** | 54.1 ± 7.3 |
| **CAT score** | 7.3 ± 5.1 |
| **DLCO, % predicted** | 72.9 ± 20.6 |
| **Comorbidities, n (%)** |  |
| Arterial hypertension | 8 (44.4) |
| Coronary or cardiovascular disease | 5 (27.8) |
| Diabetes mellitus | 3 (16.7) |
| Dyslipidemia | 3 (16.7) |
| **Medication, n (%)** |  |
| Short-acting beta agonists | 2 (11.1) |
| Long-acting beta agonists | 13 (72.2) |
| Short-acting anticholinergics | 1 (5.6) |
| Long-acting anticholinergics | 12 (66.7) |
| Inhaled corticosteroids | 10 (55.6) |
| ACE inhibitors or AT antagonists | 8 (44.4) |
| Beta blockers | 3 (16.7) |
| Calcium channel blockers | 4 (22.2) |
| Diuretics | 6 (33.3) |
| Lipid lowering therapy | 7 (38.9) |
| Antidiabetic therapy | 3 (16.7) |
| Antiplatelet aggregation therapy | 7 (38.9) |
| Antidepressants | 3 (16.7) |

Values are presented as mean ± standard deviation, or in number and percent. BMI, body mass index; FEV<sub>1</sub>, forced expiratory volume in one second; FVC, forced vital capacity. CAT, COPD assessment test; DLCO, diffusing capacity of the lung for carbon monoxide; ACE, angiotensin-converting enzyme; AT, angiotensin.

### (0) Resting condition under quiet room air breathing

The cardio- and cerebrovascular outcomes at rest are displayed in **Table 2**. At 490 m, patients were mildly hypoxemic but otherwise normocapnic and normotensive. Mean sMCAv was 48.4 ± 2.9 cm^−1^ with cerebrovascular conductance and resistance index of 0.5 ± 0 cm s^−1^ mmHg^−1^ and 2.2 ± 0.3 mmHg cm^−1^ s, respectively, and a mean CTO of 60.5 ± 1.8%.

**Table 2.** Cardio- and cerebrovascular condition(*0*) at rest breathing room air.

|  | **490 m** | **2048 m – after placebo** | **2048 m – after NOT** | **Treatment effect, NOT vs. placebo (95% CI)** | **P-value** | **2048 m – combined NOT and placebo** |
| --- | --- | --- | --- | --- | --- | --- |
| **SpO<sub>2</sub>, %** | 93.7 ± 0.4 | 90.3 ± 0.4 * | 90.1 ± 0.4 * | -0.2 (-1.1 to 0.6) | 0.613 | 90.2 ± 0.3 * |
| **PetCO<sub>2</sub>, mmHg** | 34.1 ± 1.3 | 34.4 ± 1.3 | 35.5 ± 1.3 | 1.1 (-1.4 to 3.6) | 0.395 | 35.0 ± 1.1 |
| **PtcCO<sub>2</sub>, mmHg** | 36.4 ± 1.2 | 38.1 ± 1.2 | 38.9 ± 1.2 * | 0.8 (-1.4 to 2.9) | 0.492 | 38.5 ± 1.1 * |
| **Systolic BP, mmHg** | 131 ± 4 | 135 ± 4 | 132 ± 4 | -3 (-12 to 6) | 0.502 | 134 ± 4 |
| **Diastolic BP, mmHg** | 76 ± 3 | 78 ± 3 | 74 ± 3 | -4 (-10 to 2) | 0.155 | 76 ± 2 |
| **MAP, mmHg** | 94 ± 3 | 97 ± 3 | 94 ± 3 | -4 (-10 to 2) | 0.225 | 95 ± 2 |
| **Heart rate, bpm** | 72 ± 2 | 79 ± 2 * | 78 ± 2 * | -1 (-5 to 3) | 0.566 | 78 ± 2 * |
| **sMCAv, cm/s** | 48.4 ± 2.9 | 43.3 ± 2.9 | 44.1 ± 2.9 | 0.8 (-5.9 to 7.6) | 0.810 | 43.7 ± 2 |
| **CVCi, cm/s\*mmHg** | 0.5 ± 0 | 0.5 ± 0 | 0.5 ± 0 | 0 (0 to 0.1) | 0.417 | 0.5 ± 0 |
| **CVRI, mmHg/cm\* s** | 2.2 ± 0.3 | 2.4 ± 0.3 | 2.2 ± 0.3 | -0.2 (-0.7 to 0.4) | 0.497 | 2.3 ± 0.2 |
| **CTO, %** | 60.5 ± 1.8 | 58.7 ± 1.7 | 59.0 ± 1.7 | 0.4 (-2.6 to 3.4) | 0.806 | 58.9 ± 1.6 |

Measured parameters are presented as mean ± SE, and the treatment effect as mean difference (95% CI). NOT, nocturnal oxygen therapy; SpO<sub>2</sub>, arterial oxygenation; PetCO<sub>2</sub>, end-tidal partial pressure of carbon dioxide assessed by capnography; PtcCO<sub>2</sub>, trans-cutaneous partial pressure of CO<sub>2</sub>; BP, blood pressure; MAP, mean arterial pressure measured by finger clamp technique; HR, heart rate; sMCAv, middle cerebral artery peak systolic blood flow velocity measured by transcranial Doppler ultrasound; CVCi, cerebrovascular conductance index; CVRI, cerebrovascular resistance index. CTO, cerebral tissue oxygenation measured by near-infrared spectroscopy.

At 2048 m, patients became more hypoxemic and had a higher heart rate compared to 490 m (P<0.05 both comparisons). At 2048, mean sMCAv was 43.3 ± 2.9 cm^−1^ in patients who had received placebo and 44.1 ± 2.9 cm^−1^ in patients who had received NOT, P = 0.810. The mean CVCi, CVRi and MAP were not significantly different between the NOT and placebo intervention (**Table 2**).

### Breathing maneuvers

In **Figure 2** and in **Table 3** changes of measured and calculated parameters during the conducted breathing maneuvers; *(i)* quiet oxygen breathing (hyperoxia), *(ii)* hyperventilating ambient air (hypocapnia), and *(iii)* hyperventilating oxygen (hyperoxia and hypocapnia), are summarized. The breathing maneuvers at both altitudes (490 m, 2048 m – placebo, and 2048 m – NOT) showed consistent changes in cerebral blood flow and oxygenation compared to pre-test normal breathing at the corresponding location. These changes at 2048 m were independent of the applied nocturnal intervention (NOT or placebo, **Supplemental tables S1 to S3**). Based on these observations, the data from the 2048 m – placebo and 2048 m – NOT intervention were combined and analyzed together to assess effects of breathing maneuvers on cerebral blood flow in COPD at 2048 m versus 490 m.

**Figure 2.**
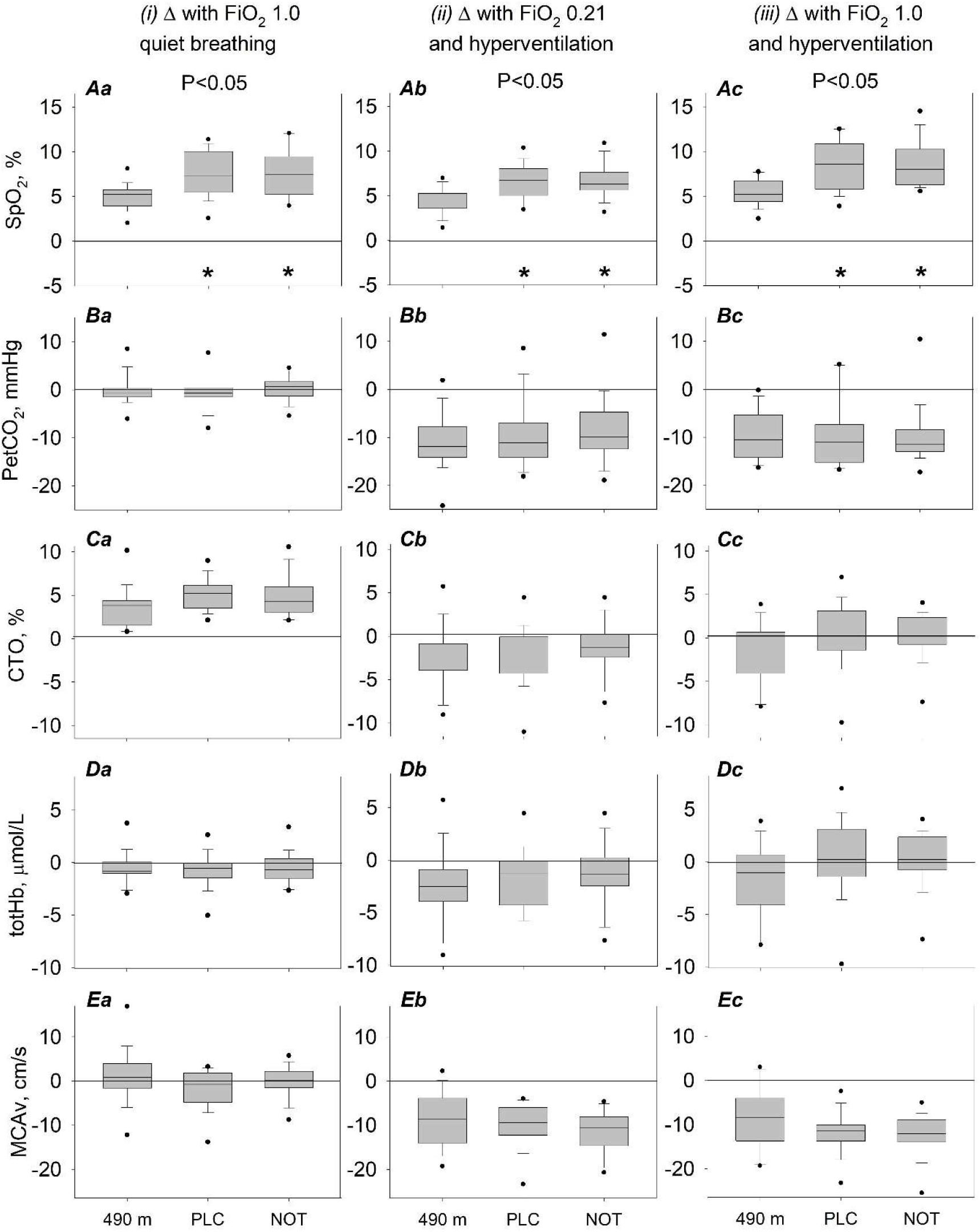
Effects of breathing maneuvers on physiological outcomes. Changes from the pre-maneuver setting are represented as bar graphs and displayed in medians (horizontal line), quartiles (boxes), 10th and 90th percentiles (whiskers) and individual values (dots) beyond this range. FiO_2_, fraction of inhaled oxygen during the breathing maneuvers; PLC, placebo at night; NOT, nocturnal oxygen therapy; SpO_2_, arterial oxygen saturation measured by oximetry; EtCO_2_, end-tidal pressure of carbon dioxide assessed by capnography; CTO, cerebral tissue oxygenation, and totHb, total hemoglobin concentration, measured by near-infrared-spectroscopy; sMCAv, middle cerebral artery peak systolic blood flow velocity measured by transcranial Doppler ultrasound. # P<0.05 vs. PLC (treatment effect); * P<0.05 vs. 490 m (altitude effect).

**Table 3.** Cardio- and cerebrovascular responsiveness to changing blood gases.

| *a)* Breathing maneuvers at 490 m |  |  |  |
| --- | --- | --- | --- |
|  | *(i) Oxygen at rest* | *(ii) Room air HV* | *(iii) Oxygen HV* |
| SpO<sub>2</sub>, % | +5.0 (4.1 to 5.9) <sup>¶</sup> | +4.3 (3.5 to 5.2) <sup>¶</sup> | +5.4 (4.5 to 6.3) <sup>¶</sup> |
| PetCO<sub>2</sub>, mmHg | 0 (-2.6 to 2.5) | -10.8 (-13.4 to -8.3) <sup>¶</sup> | -9.8 (-12.4 to -7.3) <sup>¶</sup> |
| Systolic BP, mmHg | -2 (-11 to 7) | -5 (-14 to -4) | -9 (-18 to 0) <sup>¶</sup> |
| Diastolic BP, mmHg | 0 (-6 to 6) | -1 (-7 to 4) | -0.8 (-7 to 5) |
| MAP, mmHg | -1 (-7 to 5) | -3 (-9 to 4) | -4 (-10 to 3) |
| Heart rate, bpm | -4 (-7 to -1) <sup>¶</sup> | +5 (2 to 7) <sup>¶</sup> | +4 (1 to 7) <sup>¶</sup> |
| sMCAv, cm/s | +1.0 (-5.8 to 7.8) | -8.6 (-15.4 to -1.9) <sup>¶</sup> | -8.9 (-15.6 to -2.1) <sup>¶</sup> |
| CVCi, cm/s/mmHg | 0 (-0.1 to 0.1) | -0.1 (-0.2 to 0) <sup>¶</sup> | -0.1 (-0.2 to 0) <sup>¶</sup> |
| CVRi, mmHg/cm/s | -0.1 (-0.7 to 0.5) | +0.6 (0 to 1.1) <sup>¶</sup> | +0.6 (0 to 1.2) <sup>¶</sup> |
| CTO, % | +3.6 (0.5 to 6.7) <sup>¶</sup> | -2.4 (-5.5 to 0.7) | -1.7 (-4.8 to 1.4) |
| totHb | -0.6 (-1.4 to 0.3) | +0.2 (-0.7 to 1.0) | -0.7 (-1.6 to 0.1) |
| O<sub>2</sub>Hb | +1.4 (0.6 to 2.3) <sup>¶</sup> | +0.3 (-0.6 to 1.1) | +0.5 (-0.3 to 1.4) |
| HHb | -1.9 (-2.4 to -1.5) <sup>¶</sup> | -0.1 (-0.5 to 0.3) | -1.2 (-1.7 to -0.8) <sup>¶</sup> |
| ΔsMCAv / ΔPetCO<sub>2</sub> | 0.5 ± 0.7 | 0.7 ± 0.7 | 0.7 ± 0.7 |
| ΔsMCAv / ΔSpO<sub>2</sub> | 0.2 ± 0.2 | -2.0 ± 0.2 | -1.7 ± 0.2 |
| *b)* Breathing maneuvers at 2048 m – combined groups NOT and placebo<sup>1</sup> |  |  |  |
|  | *(i) Oxygen at rest* | *(ii) Room air HV* | *(iii) Oxygen HV* |
| SpO<sub>2</sub>, % | +7.5 (6.9 to 8.1) <sup>¶</sup> * | +6.6 (6.0 to 7.3) <sup>¶</sup> * | +8.6 (8.0 to 9.2) <sup>¶</sup> * |
| PetCO<sub>2</sub>, mmHg | -0.3 (-2.1 to 1.5) | -9.1 (-10.9 to -7.3) <sup>¶</sup> | -9.7 (-11.5 to -7.9) <sup>¶</sup> |
| Systolic BP, mmHg | +0 (-6 to 7) | -5 (-11 to 2) | -6 (-12 to 1) |
| Diastolic BP, mmHg | +1 (-3 to 5) | -1 (-6 to 3) | 0 (-5 to 4) |
| MAP, mmHg | 0 (-4 to 5) | -3 (-7 to 2) | -2 (-7 to 2) |
| Heart rate, bpm | -4 (-7 to -1) <sup>¶</sup> | +5 (2 to 7) <sup>¶</sup> | +4 (2 to 7) <sup>¶</sup> |
| sMCAv, cm/s | -0.9 (-5.7 to 3.9) | -10.7 (-15.5 to -5.9) <sup>¶</sup> | -11.9 (-16.7 to -7.1) <sup>¶</sup> |
| CVCi, cm/s/mmHg | 0 (-0.1 to 0) | -0.1 (-0.2 to 0) <sup>¶</sup> | -0.1 (-0.2 to -0.1) <sup>¶</sup> |
| CVRi, mmHg/cm/s | +0.1 (-0.3 to 0.5) | +0.7 (0.3 to 1.1) <sup>¶</sup> | +0.9 (0.5 to 1.3) <sup>¶</sup> |
| CTO, % | +5.0 (2.8 to 7.1) <sup>¶</sup> | -1.6 (-3.7 to 0.5) | +0.3 (-1.8 to 2.4) |
| totHb | -0.7 (-1.2 to -0.1) <sup>¶</sup> | -0.4 (-1.0 to 0.1) | -0.6 (-1.2 to 0) <sup>¶</sup> |
| O<sub>2</sub>Hb | +2.1 (1.5 to 2.7) <sup>¶</sup> | +0.6 (0 to 1.2) <sup>¶</sup> | +1.6 (1.0 to 2.2) <sup>¶</sup> * |
| HHb | -2.8 (-3.1 to -2.5) <sup>¶</sup> * | -1.0 (-1.3 to -0.8) <sup>¶</sup> * | -2.2 (-2.5 to -1.9) <sup>¶</sup> * |
| ΔsMCAv / ΔPetCO<sub>2</sub> | 0.3 ± 0.5 | 1.1 ± 0.5 | 0.9 ± 0.5 |
| ΔsMCAv / ΔSpO<sub>2</sub> | -0.1 ± 0.2 | -1.7 ± 0.2 | -1.5 ± 0.2 |

<sup>1</sup>As no difference between NOT and placebo application was found on the outcomes during the displayed breathing maneuvers at 2048 m, the two intervention groups were combined. Detailed representation of the individual breathing maneuvers is found in the supplemental tables S1-S3. Changes from baseline (breathing room air at rest at corresponding location) are presented as mean (95% CI) for measured parameters. Calculated parameters are presented in absolute values as mean ± SE. NOT, nocturnal oxygen therapy; HV, forced hyperventilation; SpO<sub>2</sub>, arterial oxygen saturation; PetCO<sub>2</sub>, end-tidal partial pressure of carbon dioxide assessed by capnography; MAP, mean arterial pressure measured by finger clamp technique; sMCAv, middle cerebral artery peak systolic blood flow velocity measured by transcranial Doppler ultrasound; CVCi, cerebrovascular conductance index; CVRi, cerebrovascular resistance index. CTO, cerebral tissue oxygenation; totHb, total hemoglobin; O<sub>2</sub>Hb, oxygenated hemoglobin; HHb, deoxygenated hemoglobin, measured by near-infrared spectroscopy. <sup>¶</sup> P<0.05 vs. pre-test setting at corresponding location (breathing maneuver effect); \* P<0.05 vs. 490m (altitude effect).

#### (i) Quiet oxygen breathing (FiO_2_ 1.0)

At 490 m, oxygen breathing significantly increased SpO_2_, CTO and O_2_Hb, and decreased heart rate and HHb (**Table 3**). No changes were found in BP, totHb, CVCi, CVRi or sMCAv compared to quiet room air breathing. At 2048 m, totHb was decreased compared to 490 m. Also, SpO_2_ increased and HHb decreased significantly more compared to oxygen breathing at 490 m. Other cardiac and cerebrovascular parameters changed similarly.

#### (ii) Hyperventilating under room air (FiO_2_ 0.21)

At 490 m, hyperventilation under room air significantly increased SpO_2_ and heart rate, and decreased PetCO_2_ (**Table 3**). Also, CVCi increased and CVRi decreased, leading to a significant fall in mean sMCAv. BP, CTO and other parameters measured by NIRS, were not altered compared to quiet room air. At 2048 m, SpO_2_ increased and additionally HHb decreased significantly more compared to room air hyperventilation at 490 m. All other cardiac and cerebrovascular parameters changed similarly at altitude.

#### (iii) Hyperventilating under oxygen breathing (FiO_2_ 1.0)

At 490 m, hyperventilation under oxygen breathing significantly increased SpO_2_ and heart rate and decreased PetCO_2_, systolic BP and HHb (Table 3). CVCi significantly decreased and CVRi increased leading to a fall in sMCAv. No changes however were found in CTO, totHb and O2Hb compared to quiet room air. At 2048 m, SpO_2_ increased and HHb decreased significantly more compared to oxygen hyperventilation at 490 m. All other cardiac and cerebrovascular parameters changed similarly at altitude, except that systolic BP remained the same.

#### (iv) Head-up tilt manoeuvre

Same as in the analysis of the breathing maneuvers, the head-up tilt maneuver showed consistent changes in blood pressure and sMCAv drops between the placebo and NOT intervention at 2048 m (**Figure 3, Supplemental table S4**). At 490 m, the supine-to-60° tilt significantly lowered systolic and mean BP (**Table 4**). CVCi decreased and CVRi increased leading to a significant fall in sMCAv. At 2048 m, systolic, diastolic and mean arterial BP significantly decreased. CVCi, CVRi and sMCAv were similarly influenced by the maneuver at altitude compared to 490 m. Indices of cerebrovascular responsiveness to changes of blood pressure (ΔsMCAv/ΔMAP, %ΔsMCAv/ΔMAP and %ΔsMCAv/%ΔMAP) were significantly reduced at 2048 m compared to 490 m.

**Figure 3.**
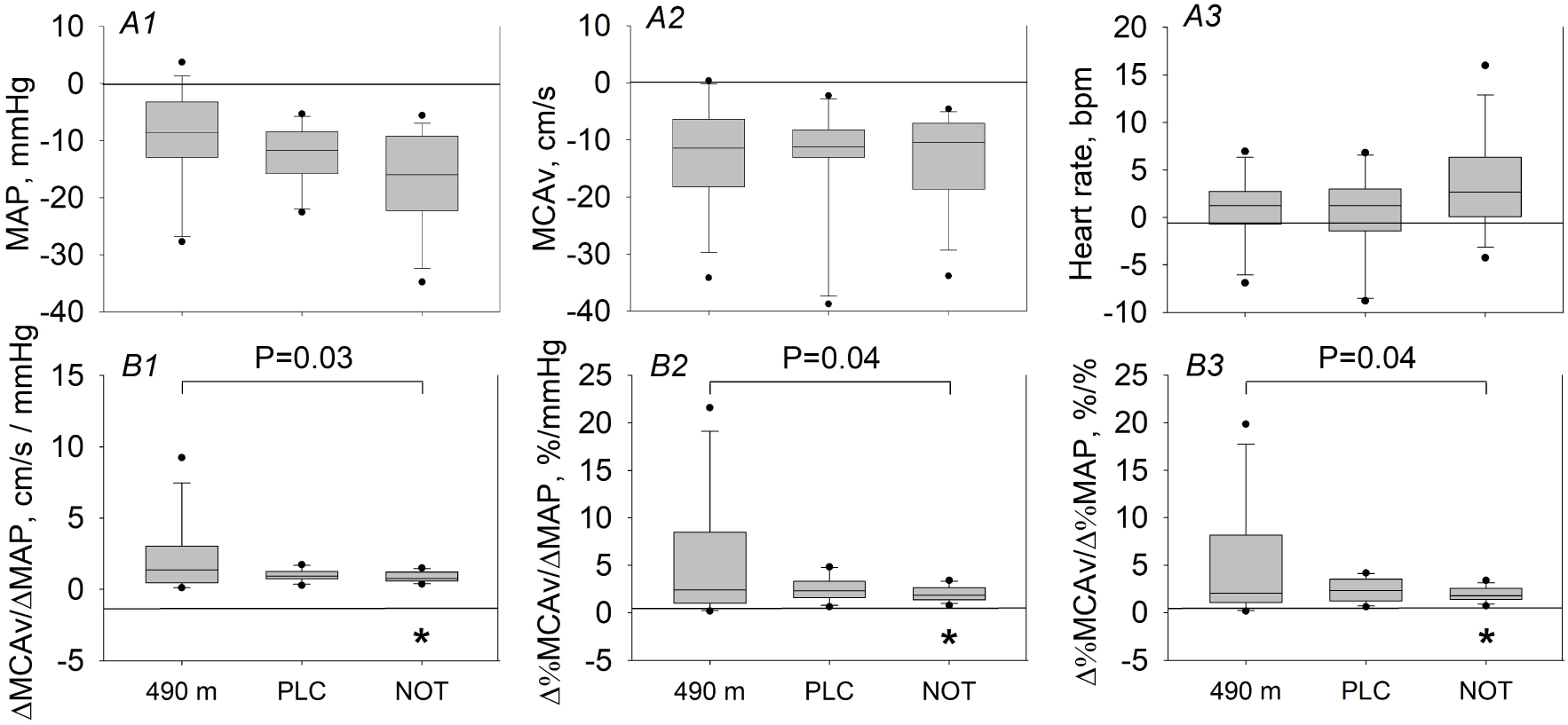
Supine-to-60° head-up tilt. Changes from the pre-maneuver setting (*A1-3*) and absolute values following the tilt maneuver (*B1-3*) are represented as bar graphs and displayed in medians (horizontal line), quartiles (boxes), 10th and 90th percentiles (whiskers) and individual values (dots) beyond this range. PLC, placebo at night; NOT, nocturnal oxygen therapy; MAP, mean arterial pressure; MCAv, middle cerebral artery peak blood flow velocity measured by transcranial Doppler ultrasound; HR, heart rate. # P<0.05 vs. PLC (treatment effect); * P<0.05 vs. 490 m (altitude effect).

**Table 4.**
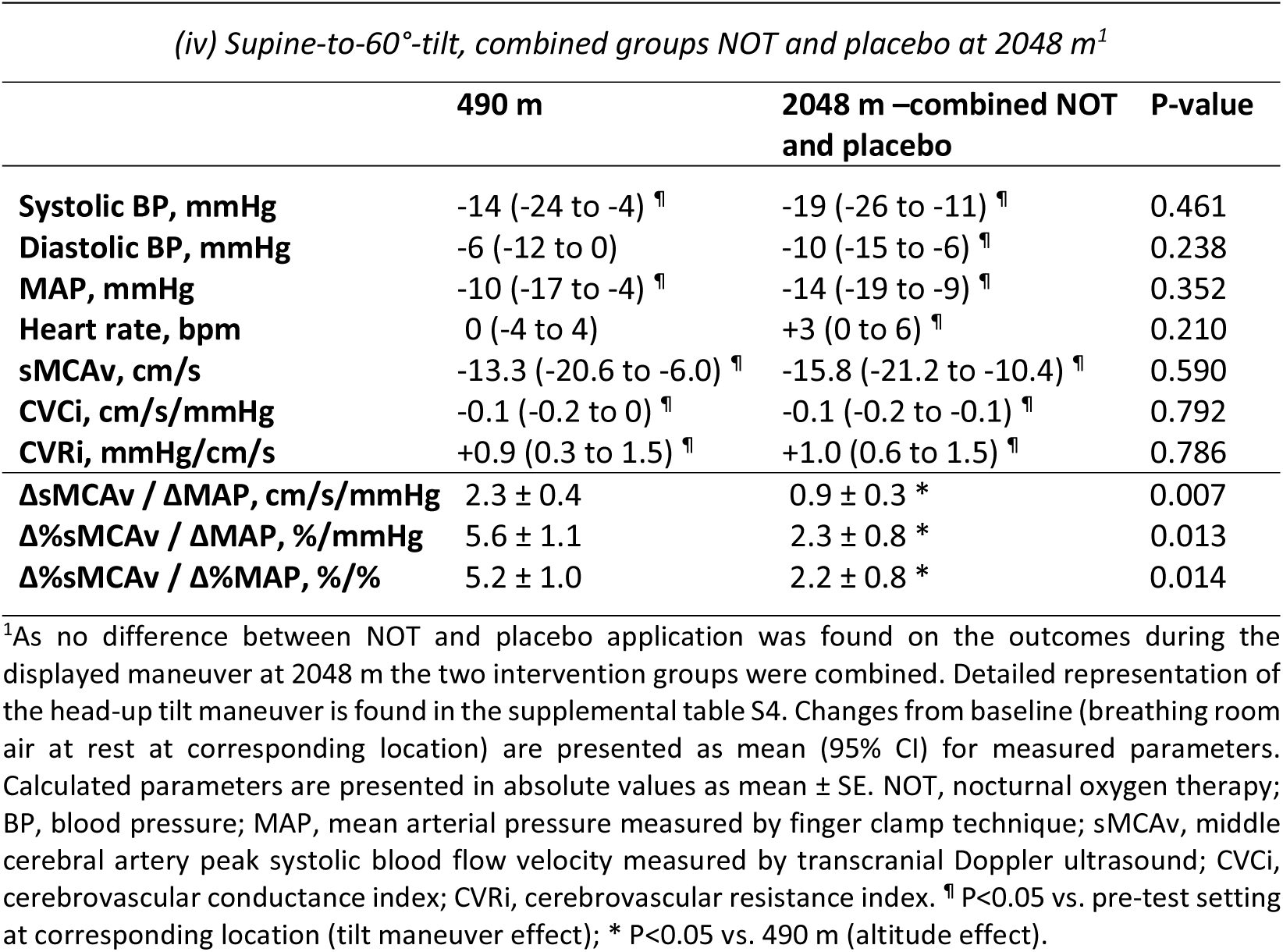
Cardio- and cerebrovascular responsiveness to systemic hypotension.

| *(iv) Supine-to-60°-tilt, combined groups NOT and placebo at 2048 m<sup>1</sup>* |  |  |  |
| --- | --- | --- | --- |
|  | **490 m** | **2048 m –combined NOT and placebo** | **P-value** |
| **Systolic BP, mmHg** | -14 (-24 to -4) <sup>¶</sup> | -19 (-26 to -11) <sup>¶</sup> | 0.461 |
| **Diastolic BP, mmHg** | -6 (-12 to 0) | -10 (-15 to -6) <sup>¶</sup> | 0.238 |
| **MAP, mmHg** | -10 (-17 to -4) <sup>¶</sup> | -14 (-19 to -9) <sup>¶</sup> | 0.352 |
| **Heart rate, bpm** | 0 (-4 to 4) | +3 (0 to 6) <sup>¶</sup> | 0.210 |
| **sMCAv, cm/s** | -13.3 (-20.6 to -6.0) <sup>¶</sup> | -15.8 (-21.2 to -10.4) <sup>¶</sup> | 0.590 |
| **CVCi, cm/s/mmHg** | -0.1 (-0.2 to 0) <sup>¶</sup> | -0.1 (-0.2 to -0.1) <sup>¶</sup> | 0.792 |
| **CVRI, mmHg/cm/s** | +0.9 (0.3 to 1.5) <sup>¶</sup> | +1.0 (0.6 to 1.5) <sup>¶</sup> | 0.786 |
| **ΔsMCAv / ΔMAP, cm/s/mmHg** | 2.3 ± 0.4 | 0.9 ± 0.3 * | 0.007 |
| **Δ%sMCAv / ΔMAP, %/mmHg** | 5.6 ± 1.1 | 2.3 ± 0.8 * | 0.013 |
| **Δ%sMCAv / Δ%MAP, %/%** | 5.2 ± 1.0 | 2.2 ± 0.8 * | 0.014 |

<sup>1</sup>As no difference between NOT and placebo application was found on the outcomes during the displayed maneuver at 2048 m the two intervention groups were combined. Detailed representation of the head-up tilt maneuver is found in the supplemental table S4. Changes from baseline (breathing room air at rest at corresponding location) are presented as mean (95% CI) for measured parameters. Calculated parameters are presented in absolute values as mean ± SE. NOT, nocturnal oxygen therapy; BP, blood pressure; MAP, mean arterial pressure measured by finger clamp technique; sMCAv, middle cerebral artery peak systolic blood flow velocity measured by transcranial Doppler ultrasound; CVCi, cerebrovascular conductance index; CVRI, cerebrovascular resistance index. <sup>¶</sup> P<0.05 vs. pre-test setting at corresponding location (tilt maneuver effect); \* P<0.05 vs. 490 m (altitude effect).

## DISCUSSION

In this randomized, crossover trial we showed that an ascent to 2048 m was tolerated by the majority of patients with moderate to severe COPD (3). At 2048 m, patients suffered from daytime hypoxemia and elevated heart rate, however, cerebrovascular determinants, i.e. CTO or sMCAv, remained unchanged compared to 490 m and were not altered the next day whether patients applied NOT or placebo during the night at altitude. Moreover, cerebrovascular responsiveness (ΔsMCAv/ΔMAP) to orthostatic hypotension was not deteriorated. These findings suggest that cerebrovascular homeostasis while at rest are maintained at moderate altitude in COPD. Correspondingly, NOT improved nocturnal oxygenation and breathing patterns compared to placebo (3) but did not modify determinants of cerebrovascular homeostasis at rest.

It is established that upon arrival at high altitude (>2500 m) CBF is acutely increased in healthy humans and remains elevated during the first few days at altitude (15). With the progression of acclimatization, CBF gradually decreases to pre-exposure values (15). The cerebrovascular response to poikilocapnic hypoxia at altitude is subdue to the cerebral vasodilation caused by hypoxemia, and the counterbalancing vasoconstriction as an effect of hypocapnia. With the increase of CBF due to hypoxemia, oxygen delivery and metabolic rate in the brain can be maintained on a relatively constant level. This was indirectly confirmed in the current study at moderate altitude, where CTO remained stable. These findings stay in contrast with the observed nocturnal cerebral hypoxia in these patients while sleeping (3) suggesting that cerebral protection might be sleep/wakefulness state and activity dependent at moderate altitude in COPD. In the same patients as in the current study, Tan Lu et al. reported that under placebo treatment at 2048 m, COPD patients had significantly lower nocturnal CTO values and elevated intermittent cerebral hypoxic events (related to sleep-disordered breathing) compared to 490 m. These findings indicate that insufficient cerebral autoregulatory compensation at night was completely eliminated with NOT at 2048 m. Lower nocturnal cerebral protection has been suggested to be related to NREM sleep since cerebral autoregulation has been proposed to be inactive during NREM compared to awake condition in COPD; and was previously observed at 2590 m (16–18). Apart of investigating quiet resting conditions in COPD at 2048 m, Gutweniger et al. investigated submaximal exercise performance and the related exercise-limiting factors at 2048 m compared to 490 m (9). She revealed that the lower endurance time at 2048 m compared to 490 m during a constant-work rate exercise testing at 60% of maximum work capacity was related, among other factors, to exercise-induced cerebral hypoxia. Similar to the current study, NOT showed no beneficial effect on next-day exercise performance. In summary, previous and current findings reveal that NREM sleep stage and cardiopulmonary challenge with exercise are capable of inducing cerebral hypoxia – but not quiet resting conditions, while staying at moderate altitude with COPD.

In the current study, we further investigated the cerebrovascular responsiveness to alterations of PaO_2_, PaCO_2_ and blood pressure using breathing maneuvers and a head-up tilt test. With the voluntary hyperventilation maneuvers under ambient air and hyperoxia (**Table 3**), we observed similar decrements in PetCO_2_ at both altitudes but higher increments in SpO_2_ without any differences in changes in CTO and in the cerebrovascular response to changes in arterial blood gases. This was confirmed by unchanged values of ΔsMCAv/ΔPetCO_2_ or ΔsMCAv/ΔSpO_2_ at 2048 m compared to 490 m. When challenging the cerebrovascular reactivity during head-up tilting, we observed similar decrements in blood pressure and sMCAv at 2048 m compared to 490 m. The significantly lower surrogate values for cerebrovascular reactivity (ΔsMCAv/ΔMAP) (**Table 4**) suggests even tighter and therefore better cerebrovascular reactivity in response to blood pressure changes at 2048 m compared to 490 m in COPD patients. These findings are in line with the overall findings of this study, suggesting that the cerebrovascular homeostasis and reactivity under resting conditions is maintained at moderate altitude in COPD. At altitude, no further COPD studies are available to compare the current results. At low altitude, Hoiland et al. (19) found the cerebral vasculature in chronic hypoxemic COPD of moderate to very severe grades (FEV_1_ = 33 %, SpO_2_ = 91 %) to be insensitive to oxygen, as no constriction in response to oxygen supplementation was observed. This circumstance may be viewed from a positive side (20) as it leads to elevated cerebral oxygen delivery and neurovascular coupling in those patients. These reported findings were confirmed in our study (**Table 3**). However, when COPD patients were more hypoxemic at 2048 m, we observed with hyperoxic gas breathing a significant decrease in totHb, a surrogate for total blood volume and cerebrovascular vasoconstriction. However, sMCAv remained unchanged despite a decrement in totHb.

### Limitations

The employed non-invasive techniques to assess cerebrovascular characteristics, TCD and NIRS, estimate surrogates of CBF and cerebral blood volume based on the assumption that the diameter of the insonated MCA remains unchanged. This assumption has been shown to be correct up to altitudes of >4000 – 5000 m (21), suggesting that no change in MCA diameter in our COPD patients at 2048 m can be expected. There needs to be noted that any influence on vascular tone and diameter via sympathetic activity influences flow and has not been quantified and corrected. Furthermore, the measured values during the orthostatic challenge have not been corrected to PaCO_2_.

## Conclusion

Based on these findings from a randomized clinical trial, we conclude that an ascent to moderate altitude does not impair resting cerebrovascular function in patients with moderate to severe COPD and may entrain even a momentary improvement in cerebrovascular reactivity to blood pressure changes on the first days at moderate altitude. Although, cerebrovascular homeostasis and reactivity at moderate altitude remain unaffected by NOT, NOT had several other beneficial effects on nocturnal indices and the incidence of altitude-related adverse health effects (3).

## Data Availability

All data produced in the present study are available upon reasonable request to the authors.

## Notes

**Funding** The study was supported by the Swiss National Science Foundation (143875) and Lunge Zurich. Siemens Health Engineers provided some equipment for the study.

### Competing Interest Statement

The authors have declared no competing interest.

### Clinical Trial

NCT02150590

### Funding Statement

The study was supported by the Swiss National Science Foundation (143875) and Lunge Zurich. Siemens Health Engineers provided some equipment for the study.

### Author Declarations

Ethics committee of the University Hospital of Zurich gave ethical approval for this work (EK-2013-0088).

