## Supplemental Material for "Effect of moderate altitude with and without nocturnal oxygen therapy on the next-day cerebrovascular function in patients with chronic obstructive pulmonary disease – A randomized, cross-over trial at 2048 m"

**Running head:** Oxygen therapy in COPD at altitude

Dominic Gilliand[1], Tsogyal D. Latshang[1], Sayaka S. Aeschbacher[1], Fabienne Huber[1], Deborah Flueck[1], Mona Lichtblau[1], Stefanie Ulrich[1], Elisabeth D. Hasler[1], Philipp M. Scheiwiller[1], Julian Müller[1], Silvia Ulrich[1], Konrad E. Bloch[1], Michael Furian[1,2]

[1]Department of Respiratory Medicine, University Hospital of Zurich, Zurich, Switzerland

[2]Research Department, Swiss University of Traditional Chinese Medicine, Bad Zurzach, Switzerland

**Funding**

The study was supported by the Swiss National Science Foundation (143875) and Lunge Zurich.

Siemens Health Engineers provided some equipment for the study.

**Correspondence**

Michael Furian, Dr. sc. ETH
University Hospital Zurich
Pulmonology Department
Raemistrasse 100
8092 Zurich


**Table S1. Effects of applying NOT vs. placebo on next day cardio- and cerebrovascular indices in patients with COPD at altitude –response to *i)* breathing oxygen (hyperoxia)**

| | 490 m | 2048 m - placebo | 2048 m - NOT | Treatment effect | P value |
|---|---|---|---|---|---|
| **$SpO_2$, %** | +5.0 (4.1 to 5.9) ¶ | +7.5 (6.6 to 8.3) ¶* | +7.6 (6.7 to 8.5) ¶* | +0.1 (-1.1 to 1.3) | 0.862 |
| **$PetCO_2$, mmHg** | 0 (-2.6 to 2.5) | -0.7 (-3.2 to 1.8) | +0.2 (-2.3 to 2.7) | +0.9 (-2.7 to 4.4) | 0.624 |
| **Systolic BP, mmHg** | -2 (-11 to 7) | -1 (-10 to 8) | +1 (-8 to 10) | +2 (-11 to 15) | 0.745 |
| **Diastolic BP, mmHg** | 0 (-6 to 5) | +1 (-5 to 6) | 0 (-5 to 6) | 0 (-8 to 8) | 0.930 |
| **MAP, mmHg** | -1 (-7 to 5) | 0 (-6 to 6) | +1 (-5 to 7) | 0 (-8 to 9) | 0.913 |
| **Heart rate, bpm** | -4 (-7 to 0) | -4 (-8 to 0) ¶ | -4 (-8 to 0) ¶ | 0 (-5 to 5) | 0.967 |
| **sMCAv, cm/s** | +1.0 (-5.8 to 7.8) | -1.7 (-8.5 to 5.0) | 0 (-6.8 to 6.8) | +1.7 (-7.9 to 11.3) | 0.725 |
| **CVCi, cm/s/mmHg** | 0 (-0.1 to 0.1) | 0 (-0.1 to 0.1) | 0 (-0.1 to 0.1) | 0 (-0.1 to 0.1) | 0.714 |
| **CVRi, mmHg/cm/s** | -0.1 (-0.7 to 0.5) | +0.2 (-0.3 to 0.8) | 0 (-0.5 to 0.6) | -0.2 (-1.0 to 0.6) | 0.601 |
| **CTO, %** | +3.6 (0.5 to 6.7) ¶ | +5.1 (2.1 to 8.0) ¶ | +4.9 (1.9 to 7.9) ¶ | -0.2 (-4.4 to 4.1) | 0.938 |
| **totHb** | -0.6 (-1.4 to 0.3) | -0.7 (-1.6 to 0.1) | -0.6 (-1.4 to 0.2) | +0.2 (-1.0 to 1.3) | 0.767 |
| **$O_2Hb$** | +1.4 (0.6 to 2.3) ¶ | +2.0 (1.2 to 2.9) ¶ | +2.2 (1.4 to 3.0) ¶ | +0.2 (-1.0 to 1.3) | 0.799 |
| **HHb** | -1.9 (-2.4 to -1.5) ¶ | -2.8 (-3.2 to -2.4) ¶* | -2.8 (-3.2 to -2.4) ¶* | 0 (-0.6 to 0.6) | 0.999 |
| **$\Delta$sMCAv/ $\Delta PetCO_2$** | 0.5 ± 0.7 | 0.4 ± 0.7 | 0.2 ± 0.7 | -0.2 (-2.1 to 1.7) | 0.837 |
| **$\Delta$sMCAv/ $\Delta SpO_2$** | 0.2 ± 0.2 | -0.2 ± 0.2 | 0.1 ± 0.2 | +0.3 (-0.3 to 0.9) | 0.354 |

Changes from baseline (breathing room air at rest at corresponding location) are presented as mean (95% CI) for measured parameters. Calculated parameters are presented in absolute values as mean ± SE, and the treatment effect as mean difference (95% CI). NOT, nocturnal oxygen therapy; $SpO_2$, arterial oxygen saturation; $PetCO_2$, end-tidal partial pressure of carbon dioxide assessed by capnography; MAP, mean arterial pressure measured by finger clamp technique; sMCAv, middle cerebral artery peak systolic blood flow velocity measured by transcranial Doppler ultrasound; CVCi, cerebrovascular conductance index; CVRi, cerebrovascular resistance index; CTO, cerebral tissue oxygenation measured by near-infrared spectroscopy; totHb, total hemoglobin; $O_2Hb$, oxygenated hemoglobin; HHb, deoxygenated hemoglobin.

### P<0.05 NOT vs. PLC (treatment effect)

¶ P<0.05 vs. rest at corresponding location (breathing maneuver effect)

* P<0.05 vs. change at 490 m (altitude effect)

**Table S2. Effects of applying NOT vs. placebo on next day cardio- and cerebrovascular indices in patients with COPD at altitude – response *ii)* to room air hyperventilation (hypocapnia)**

| | 490 m | 2048 m - placebo | 2048 m - NOT | Treatment effect | P value |
|---|---|---|---|---|---|
| **$SpO_2$, %** | +4.4 (3.5 to 5.2) ¶ | +6.6 (5.8 to 7.5) ¶* | +6.6 (5.8 to 7.5) ¶* | 0 (-1.2 to 1.2) | 0.996 |
| **$PetCO_2$, mmHg** | -10.8 (-13.3 to -8.3) ¶ | -9.6 (-12.1 to -7.0) ¶ | -8.7 (-11.2 to -6.2) ¶ | +0.8 (-2.7 to 4.4) | 0.644 |
| **Systolic BP, mmHg** | -5 (-14 to 4) | -8 (-17 to 1) | -2 (-11 to 8) | +6 (-7 to 19) | 0.342 |
| **Diastolic BP, mmHg** | -1 (-7 to 4) | -3 (-8 to 3) | 0 (-6 to 6) | +3 (-5 to 11) | 0.488 |
| **MAP, mmHg** | -3 (-9 to 3) | -5 (-11 to 2) | -1 (-7 to 5) | +4 (-5 to 13) | 0.365 |
| **Heart rate, bpm** | +6 (3 to 10) ¶ | +4 (1 to 8) ¶ | +5 (1 to 8) ¶ | +1 (-4 to 6) | 0.807 |
| **sMCAv, cm/s** | -8.6 (-15.4 to -1.9) ¶ | -10.0 (-16.7 to -3.2) ¶ | -11.5 (-18.3 to -4.7) ¶ | -1.6 (-11.1 to 8.0) | 0.750 |
| **CVCi, cm/s/mmHg** | -0.1 (-0.2 to 0.0) ¶ | -0.1 (-0.2 to 0.0) ¶ | -0.1 (-0.2 to 0.0) ¶ | 0.0 (-0.1 to 0.1) | 0.532 |
| **CVRi, mmHg/cm/s** | +0.6 (0 to 1.1) ¶ | +0.6 (0.0 to 1.1) ¶ | +0.8 (0.3 to 1.4) ¶ | +0.2 (-0.6 to 1.0) | 0.553 |
| **CTO, %** | -2.4 (-5.5 to 0.7) | -1.8 (-4.8 to 1.2) | -1.3 (-4.3 to 1.7) | +0.5 (-3.7 to 4.7) | 0.817 |
| **totHb** | +0.2 (-0.7 to 1.0) | -0.5 (-1.3 to 0.3) | -0.4 (-1.2 to 0.4) | +0.1 (-1.1 to 1.2) | 0.867 |
| **$O_2Hb$** | +0.3 (-0.6 to 1.2) | +0.6 (-0.3 to 1.4) | +0.6 (-0.2 to 1.5) | 0.0 (-1.1 to 1.2) | 0.951 |
| **HHb** | -0.1 (-0.6 to 0.3) | -1.1 (-1.5 to -0.7) ¶* | -1.0 (-1.4 to -0.6) ¶* | 0.0 (-0.5 to 0.6) | 0.879 |
| **ΔsMCAv/ $\Delta PetCO_2$** | 0.7 ± 0.7 | 0.6 ± 0.7 | 1.6 ± 0.7 | +1.0 (-0.9 to 3.0) | 0.306 |
| **ΔsMCAv/ $\Delta SpO_2$** | -2.0 ± 0.3 | -1.6 ± 0.3 | -1.8 ± 0.3 | -0.2 (-0.8 to 0.4) | 0.593 |

Changes from baseline (breathing room air at rest at corresponding location) are presented as mean (95% CI) for measured parameters. Calculated parameters are presented in absolute values as mean ± SE, and the treatment effect as mean difference (95% CI). NOT, nocturnal oxygen therapy; $SpO_2$, arterial oxygen saturation; $PetCO_2$, end-tidal partial pressure of carbon dioxide assessed by capnography; MAP, mean arterial pressure measured by finger clamp technique; sMCAv, middle cerebral artery peak systolic blood flow velocity measured by transcranial Doppler ultrasound; CVCi, cerebrovascular conductance index; CVRi, cerebrovascular resistance index; CTO, cerebral tissue oxygenation measured by near-infrared spectroscopy; totHb, total hemoglobin; $O_2Hb$, oxygenated hemoglobin; HHb, deoxygenated hemoglobin.

### P<0.05 NOT vs. PLC (treatment effect)

¶ P<0.05 vs. rest at corresponding location (breathing maneuver effect)

* P<0.05 vs. change at 490 m (altitude effect)

**Table S3. Effects of applying NOT vs. placebo on next day cardio- and cerebrovascular indices in patients with COPD at altitude – response to *iii)* oxygen hyperventilation (hyperoxia and hypocapnia)**

| | 490 m | 2048 m - placebo | 2048 m - NOT | Treatment effect | P value |
|---|---|---|---|---|---|
| **$SpO_2$, %** | +5.4 (4.5 to 6.2) ¶ | +8.5 (7.7 to 9.4) ¶* | +8.7 (7.8 to 9.5) ¶* | +0.2 (-1.1 to 1.4) | 0.797 |
| **$PetCO_2$, mmHg** | -9.8 (-12.3 to -7.3) ¶ | -9.7 (-12.2 to 7.2) ¶ | -9.8 (-12.3 to -7.3) ¶ | -0.1 (-3.7 to 3.5) | 0.956 |
| **Systolic BP, mmHg** | -9 (-18 to 0) ¶ | -7 (-17 to 2) | -4 (-13 to 5) | +4 (-9 to 16) | 0.594 |
| **Diastolic BP, mmHg** | -1 (-6 to 5) | 0 (-6 to 5) | -1 (-6 to 5) | -1 (-8 to 7) | 0.900 |
| **MAP, mmHg** | -4 (-10 to 2) | -3 (-9 to 3) | -2 (-8 to 4) | +1 (-8 to 9) | 0.849 |
| **Heart rate, bpm** | +5 (1 to 9) ¶ | +4 (1 to 8) ¶ | +4 (0 to 8) ¶ | 0 (-5 to 5) | 0.956 |
| **MCAv, cm/s** | -8.9 (-15.6 to -2.1) ¶ | -11.7 (-18.5 to -4.9) ¶ | -12.2 (-19.0 to -5.4) ¶ | -0.5 (-10.1 to 9.1) | 0.915 |
| **CVCi, cm/s/mmHg** | -0.1 (-0.2 to 0.0) ¶ | -0.1 (-0.2 to 0.0) ¶ | -0.1 (-0.2 to 0.0) ¶ | 0 (-0.1 to 0.1) | 0.849 |
| **CVRi, mmHg/cm/s** | +0.6 (0.0 to 1.1) ¶ | +1.0 (0.4 to 1.5) ¶ | +0.8 (0.2 to 1.4) ¶ | -0.2 (-1.0 to 0.6) | 0.655 |
| **CTO, %** | -1.7 (-4.8 to 1.4) | +0.4 (-2.6 to 3.4) | +0.2 (-2.8 to 3.2) | -0.2 (-4.4 to 4.0) | 0.930 |
| **totHb** | -0.7 (-1.6 to 0.1) | -0.6 (-1.4 to 0.2) | -0.6 (-1.4 to 0.2) | 0 (-1.1 to 1.1) | 0.984 |
| **$O_2Hb$** | +0.5 (-0.3 to 1.3) | +1.6 (0.8 to 2.4) ¶ | +1.6 (0.7 to 2.4) ¶ | 0 (-1.2 to 1.1) | 0.943 |
| **HHb** | -1.2 (-1.7 to -0.8) ¶ | -2.2 (-2.6 to -1.8) ¶* | -2.2 (-2.6 to -1.8) ¶* | 0 (-0.5 to 0.6) | 0.896 |
| **ΔMCAv/ $\Delta PetCO_2$** | 0.7 ± 0.7 | 0.7 ± 0.7 | 1.1 ± 0.7 | +0.3 (-1.6 to 2.3) | 0.738 |
| **ΔMCAv/ $\Delta SpO_2$** | -1.7 ± 0.3 | -1.5 ± 0.3 | -1.5 ± 0.3 | 0.0 (-0.6 to 0.6) | 0.960 |

Changes from baseline (breathing room air at rest at corresponding location) are presented as mean (95% CI) for measured parameters. Calculated parameters are presented in absolute values as mean ± SE, and the treatment effect as mean difference (95% CI). NOT, nocturnal oxygen therapy; $SpO_2$, arterial oxygen saturation; $PetCO_2$, end-tidal partial pressure of carbon dioxide assessed by capnography; MAP, mean arterial pressure measured by finger clamp technique; MCAv, middle cerebral artery peak blood flow velocity measured by transcranial Doppler ultrasound; CVCi, cerebrovascular conductance index; CVRi, cerebrovascular resistance index; CTO, cerebral tissue oxygenation measured by near-infrared spectroscopy; totHb, total hemoglobin; $O_2Hb$, oxygenated hemoglobin; HHb, deoxygenated hemoglobin.
### P<0.05 NOT vs. PLC (treatment effect)
¶ P<0.05 vs. rest at corresponding location (breathing maneuver effect)
* P<0.05 vs. change at 490 m (altitude effect)

**Table S4. Effects of applying NOT vs. placebo on next day cardio- and cerebrovascular indices in patients with COPD at altitude – response to a i*v)* supine-to-60° head-up-tilt (orthostatic hypertension)**

| | 490 m | 2048 m - placebo | 2048 m - NOT | Treatment effect | P value |
|---|---|---|---|---|---|
| **Systolic BP, mmHg** | -14 (-24 to -4) ¶ | -17 (-28 to -7) ¶ | -19 (-29 to -9) ¶ | -2 (-17 to 12) | 0.790 |
| **Diastolic BP, mmHg** | -6 (-12 to 0) | -8 (-15 to -2) ¶ | -12 (-18 to -6) ¶ | -4 (-12 to 6) | 0.462 |
| **MAP, mmHg** | -10 (-17 to -4) ¶ | -12 (-19 to -5) ¶ | -15 (-22 to -9) ¶ | -3 (-13 to 7) | 0.540 |
| **Heart rate, bpm** | 0 (-4 to 4) | +2 (-2 to 6) | +4 (0 to 8) (0.052) | +2 (-4 to 8) | 0.523 |
| **MCAv, cm/s** | -13.3 (-20.5 to -6.0) ¶ | -16.8 (-24.7 to -9.0) ¶ | -14.9 (-22.4 to -7.5) ¶ | -1.9 (-8.9 to 12.7) | 0.605 |
| **CVCi, cm/s/mmHg** | -0.1 (-0.2 to 0) ¶ | -0.1 (-0.2 to 0) ¶ | -0.1 (-0.2 to 0) ¶ | 0 (-0.1 to 0.2) | 0.991 |
| **CVRi, mmHg/cm/s** | +0.9 (0.3 to 1.5) ¶ | +1.4 (0.7 to 2.0) ¶ | +0.7 (0.1 to 1.3) ¶ | -0.6 (-1.5 to 0.3) | 0.166 |
| **ΔMCAv/ΔMAP, cm $s^{-1}$ mmHg** | 2.3 ± 0.7 | 1.0 ± 0.1 (p=0.060) | 0.9 ± 0.1 * (p=0.034) | -0.2 (-0.5 to 0.1) | 0.310 |
| **Δ%MCAv / ΔMAP, %/mmHg** | 5.6 ± 1.7 | 2.6 ± 0.4 (p=0.090) | 2.0 ± 0.2 * (p=0.040) | -0.6 (1.4 to 0.3) | 0.194 |
| **Δ%MCAv / Δ%MAP, %/%** | 5.2 ± 1.6 | 2.5 ± 0.4 (p=0.098) | 1.9 ± 0.2 * (p=0.043) | -0.6 (-1.4 to 0.2) | 0.166 |

Changes from baseline (breathing room air at rest at corresponding location) are presented as mean (95% CI) for measured parameters. Calculated parameters are presented in absolute values as mean ± SE, and the treatment effect as mean difference (95% CI). NOT, nocturnal oxygen therapy; BP, blood pressure; MAP, mean arterial pressure measured by finger clamp technique; MCAv, middle cerebral artery peak blood flow velocity measured by transcranial Doppler ultrasound; CVCi, cerebrovascular conductance index; CVRi, cerebrovascular resistance index.

¶ P<0.05 vs. during rest at corresponding location (tilt maneuver effect)

* P<0.05 vs. 490 m (altitude effect)
